# Investigating the Influence of prolonged Stroop task on mental fatigue and the consequences on corticospinal and corticocortical excitability: A pilot study

**DOI:** 10.1101/2023.11.28.23299165

**Authors:** Abubakar Tijjani Salihu, Keith D. Hill, Maryam Zoghi, Shapour Jaberzadeh

**Affiliations:** Monash Neuromodulation Research Unit, Department of Physiotherapy, School of Primary and Allied Health Care, Faculty of Medicine, Nursing and Health Science, Monash University, Melbourne, Australia; Rehabilitation, Ageing and Independent Living (RAIL) Research Centre, School of Primary and Allied Health Care, Monash University, Frankston, Australia; Discipline of Physiotherapy, Institute of Health and Wellbeing, Federation University Australia

**Keywords:** Prolonged cognitive activity, Mental fatigue, Corticospinal excitability, Cortico-cortical excitability

## Abstract

Changes in the corticospinal (CSE) and cortico-cortical (CCE) excitability of the primary motor cortex (M1) may underlie the effect of mental fatigue on physical performance. To date, research on this subject has predominantly focused on the examination of CSE, with limited exploration of effects of mental fatigue on CCE. This study aims to investigate the influence of mental fatigue induced through prolonged cognitive activity on both CSE and CCE. Fifteen healthy adults (aged 29.13±7.15 years) participated in assessments of CSE (Motor evoked potential - MEP amplitude) and CCE (Intracortical facilitation - ICF, short-interval intracortical inhibition - SICI, and long-interval intracortical inhibition - LICI) before and after a 60-minute Stroop task (experimental condition) or watching a documentary (control condition). Subjective mental fatigue was measured using the mental fatigue visual analogue scale (M-VAS), and workload associated with the tasks was assessed using the National Aeronautics and Space Administration (NASA) task load index. Objective mental fatigue was defined by the time-related decline in Stroop task performance. The study results revealed no significant differences in M-VAS, CSE and CCE between the two conditions. Stroop task performance did not exhibit significant changes over time. However, participants perceived the Stroop task to be more mentally demanding and effortful than watching the documentary (p<0.05). Further analysis of Stroop task performance at individual participants level identified two sub-groups of participants: one exhibiting deteriorating performance with time (fatigued subgroup) and the other showing improved performance (non-fatigued subgroup). Descriptively, cortical inhibition increased (reduced SICI and ICF values) from pre to post Stroop task in the fatigued subgroup, while the non-fatigued group displayed an opposite pattern. The findings suggest that mental fatigue may lead to increased cortical inhibition, highlighting the need for further investigation with a larger sample size.

## 1. Introduction

Modern society is characterized by various activities which require prolonged engagement in intense or monotonous cognitive tasks demanding sustained attention (1, 2). Many tasks including social activities such as the use of social media and video gaming, occupational activities, sports, and other day to day activities involve one form of cognitive engagement or another (3–7). These lifestyle factors lead to considerable complaints of mental fatigue in the modern society (1, 4–6, 8–12). Mental fatigue is a psychobiological state characterized by a sensation of mental tiredness or exhaustion and a decrease in cognitive performance, following a prolonged or sustain cognitive activity (13, 14). In addition to the mental fatigue caused by lifestyle factors, many chronic diseases including stroke, Multiple Sclerosis, Parkinson’s disease, as well as mental illnesses such as depression and anxiety present with mental fatigue, further contributing to the burden of this condition in the society (15–19). A distinctive feature of mental fatigue is its effect on cognitive performance, which can have serious consequences on both productivity and safety at work, as well as overall daily functioning of the affected individuals (12, 13, 20). Moreover, studies have shown that mental fatigue can also affect physical performance in activities such as whole-body endurance (aerobic performance), muscular endurance (isometric and dynamic resistance), motor skills performance and postural balance, in addition to its effect on cognitive performance (21–25). Therefore, prior cognitive exertion leading to mental fatigue may affect human physical performance during professional or recreational sports, social, occupational, or other day-to-day activities that require aerobic fitness, muscular endurance, or motor skills (21). Additionally, mental fatigue may predispose people to falls and fall-related injuries through its effect on the postural balance (26, 27).

Consequently, there is increased interest in understanding the brain mechanisms underlying the effects of mental fatigue on physical performance (28). This is of utmost importance as it will establish a critical foundation for future research aimed at investigating both pharmacological and non-pharmacological interventions to mitigate the impact of mental fatigue on physical performance. As the primary structure that generate the motor commands to the muscles for various physical tasks, changes in the excitability of the primary motor cortex (M1) may underlie the impaired motor performance that developed due to mental fatigue (28, 29). Over the years, several studies have investigated the changes in the excitability of the M1 following prolonged or sustained cognitive exertion leading to mental fatigue (28, 30–33). However, the findings of these studies are inconsistent with some studies showing an increase in corticospinal excitability (32), while others found a decrease (33) or no change (28, 30, 31). In addition to methodological differences, all the studies have primarily focused on changes in transcranial magnetic stimulation (TMS) evoked motor evoked potentials (MEPs) peak-to-peak amplitude which is an index of corticospinal excitability (CSE). The CSE is a product of the excitability of various neuronal structures and circuits including the excitatory and inhibitory circuits in the M1 (cortico-cortical excitability-CCE), the spinal motor neurons and the corticospinal tract (34). Intracortical facilitation (ICF), short interval intracortical inhibition (SICI), and long interval intracortical inhibition (LICI) are important metrics of CCE that contribute to the overall CSE (35). An elevation in the ICF parameter signifies heightened facilitation within the cortex, whereas an increase in SICI and LICI parameters suggests reduced inhibition. Both alterations can result in an increase in CCE and consequently the CSE. Conversely, a decline in ICF parameter signifies diminished cortical facilitation, whereas decreased SICI and LICI parameters indicate heightened inhibition within the cortex. Both changes can lead to a reduction in CCE and subsequently the CSE (36, 37).

As a phenomenon principally arising from cognitive task performance, effects of mental fatigue may primarily affect the cortical circuits, which may then bring about changes in other peripheral structures in a top-down manner (38, 39). Hence, examining the changes in measures of CCE within the M1 may provide better information about the potential neural mechanisms underlining the effect of mental fatigue on physical performance. This was not examined in any of the previously published studies. The primary aim of this study is to investigate the effects of prolonged cognitive activity on subjective and objective mental fatigue, and any potential effect thereafter on CSE and underlying inhibitory and facilitatory CCE mechanisms.

## 2. Methodology

### 2.1. Participants

Fifteen healthy adults (11 males, 4 females; mean age 29.13±7.15 years) took part in the study. All participants were right-handed as determined by the short form of Edinburgh Handedness Inventory (40). Exclusion criteria were having a neurological, musculoskeletal, psychiatric, visual or sleep disorder. People taking medications that could alter their level of physical or cognitive functions were similarly excluded. Written informed consent was obtained from all participants. The study was approved by the local ethics committee of Monash University, Melbourne, Australia (Project ID: 27394).

### 2.2. Study design

The study was based on a randomized, counterbalanced, cross-over design. Experiments were conducted in the Non-invasive Brain Stimulation and Neuroplasticity Laboratory, Department of Physiotherapy, Monash University, Melbourne, Australia. Participants were assigned to attend the laboratory for data collection on two separate occasions, once for the experimental session, involving mental fatigue, and once for the control session, involving watching a documentary (Figure 1). The order of these sessions was randomized for each participant. A minimum of three days elapsed between the two sessions to facilitate proper washout and mitigate any potential carryover effects.

**Figure 1.**
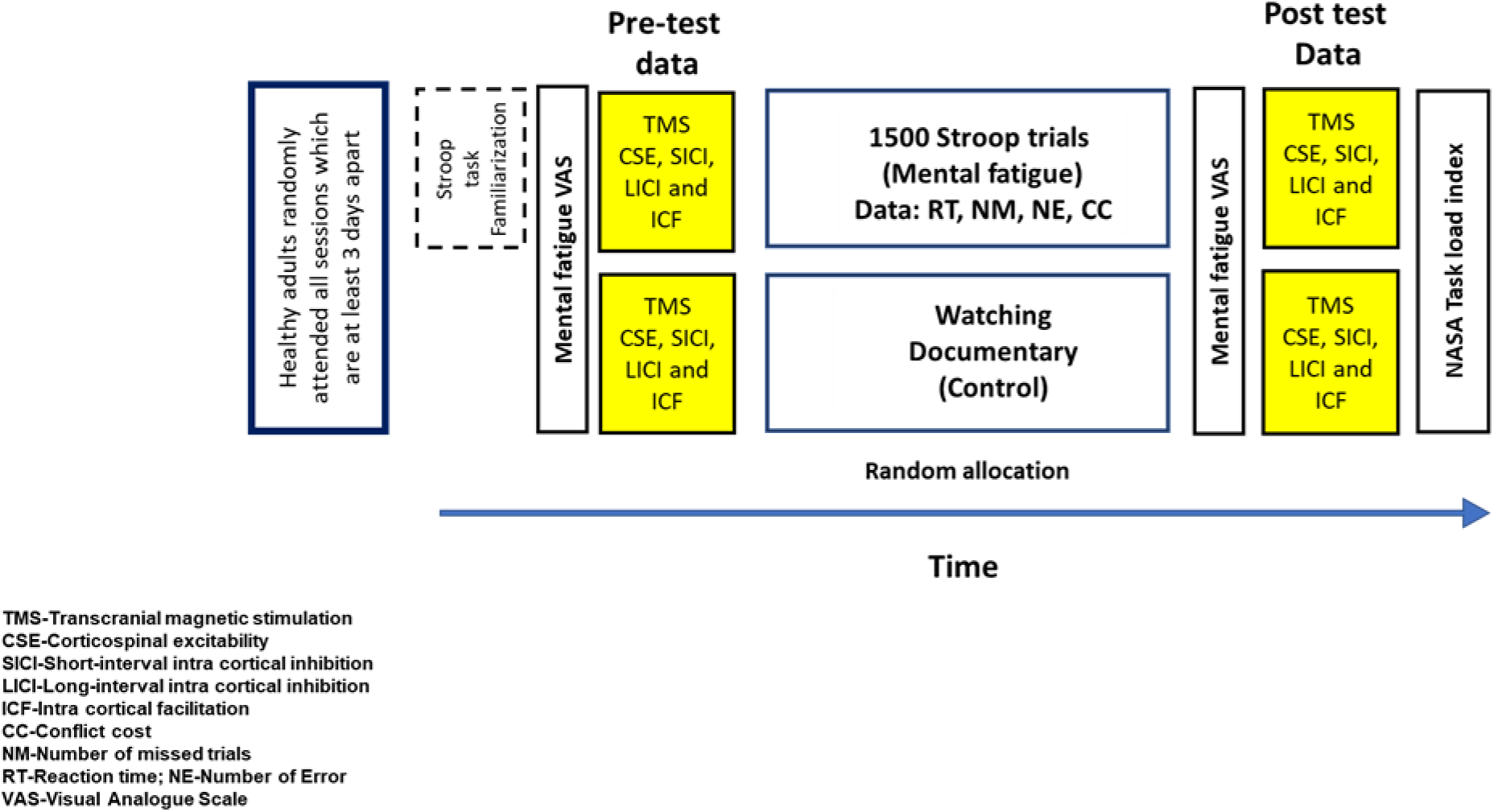
Study design

Participants were instructed to ensure adequate sleep (at least 7 h) the night before; refrain from consumption of caffeine or alcohol 12 h before; and not to engage in rigorous physical exercise 24 h before each of the sessions. To control circadian effects, both the experimental and control sessions were carried out at the same time of the day. In each session, evaluations of CSE and CCE were conducted both before and after the interventions (either the experimental or control sessions) using TMS. Before application of the TMS, participants completed a TMS safety questionnaire (41) to ensure that they were safe to undergo stimulation using this technique.

### 2.3. Experimental and Control Interventions

#### 2.3.1. Experimental Intervention

In the experimental condition, where the aim was to induce mental fatigue due to prolonged cognitive activity, the intervention involved performance of a Stroop task (42). The task contained 1500 Stroop trials, 75% of which are incongruent, while the remaining trials were congruent. The congruent and incongruent trials were presented at random, continuously without a break until the last trial, and the overall task duration was about one hour (mean task duration: 59.5 minutes; Standard deviation (SD): 3.26).

In the incongruent trial, one of four coloured words (“red,” “yellow,” “green,” or “blue”) was presented on a computer screen. The meaning of the word and the colour of the ink it was printed in were different (e.g., the word “green” printed in “red” ink), and participants were instructed to ignore the meaning of the word and respond based on the colour of the ink. The response was carried out via computer-linked foot switches, by pressing the particular foot switch labelled with the letter matching the initial letter of that colour using the lateral aspect of the right clenched fist (e.g., “R” for red or “G” for green) (43) (Figure 2). This setup was implemented to prevent activity, and potentially physical fatigue, in the target muscle for transcranial magnetic stimulation (first dorsal interossei) that might occur if the participants respond by pressing the corresponding letters on the computer keyboard using the fingers.

**Figure 2.**
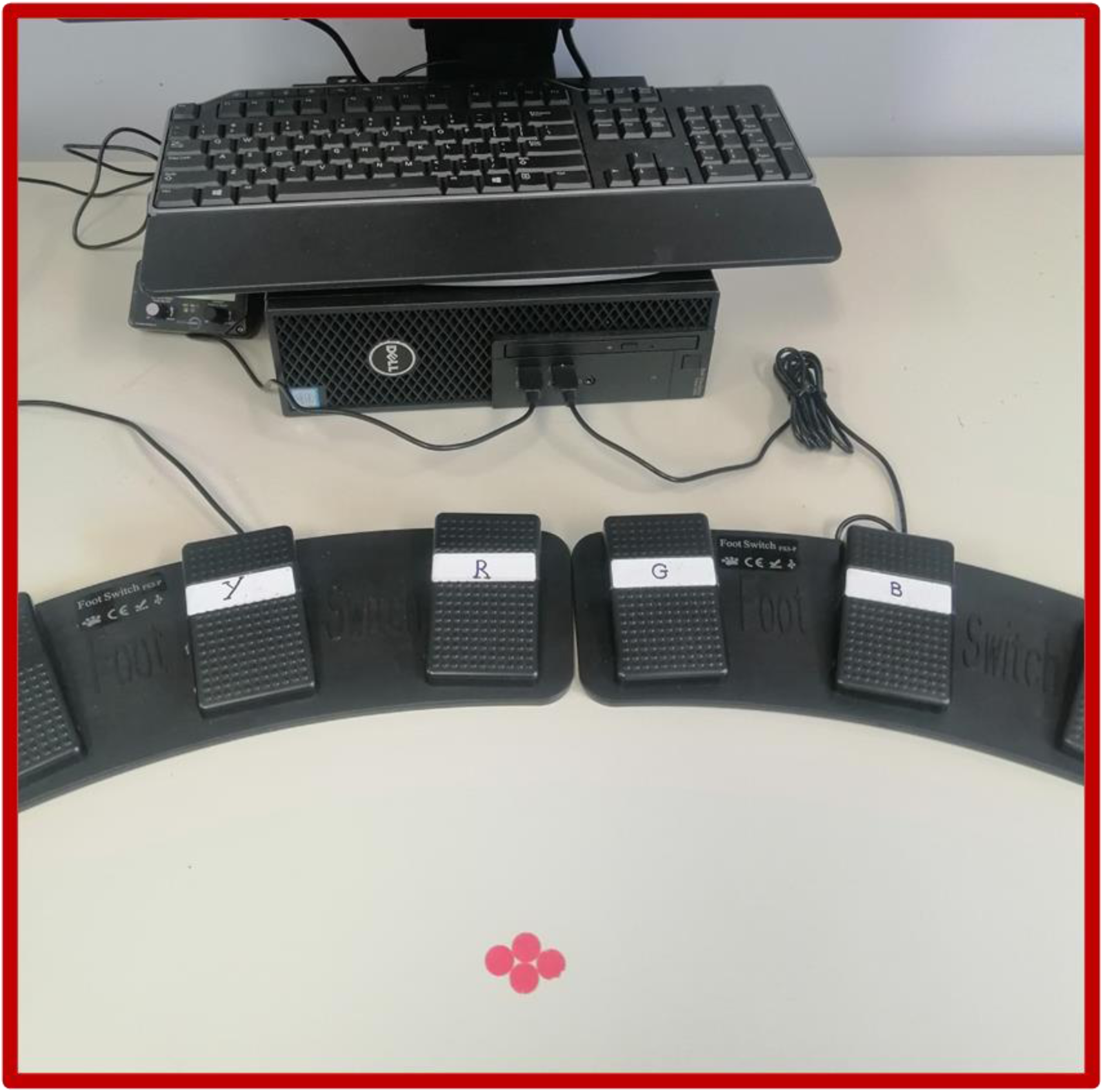
Image showing the computer-linked foot switches used for providing response during the Stroop task.

For the congruent trials, the meaning of the word and the colour of the ink it was printed in were the same, and participants responded by pressing the foot switch corresponding to the presented coloured word (43). In each trial, the coloured word remained on the computer screen for 2000 milliseconds, and if the participant couldn’t respond within this time frame, the trial was automatically recorded as a missed trial (43). Otherwise, the time between the presentation of the stimulus and the participant’s response was automatically recorded as the reaction time for that trial. Similarly, incorrect responses were automatically recorded as errors, and feedback indicating correct or incorrect responses appeared on the computer screen after each trial before the next trial was presented (43). Participants were instructed to respond as quickly and accurately as possible. Before starting the Stroop task, participants performed a practice task containing 80 Stroop trials to familiarize themselves with the task. The Stroop task was conducted using the Psytoolkit platform, a free-to-use toolkit for demonstrating, programming, and running cognitive experiments (44, 45).

#### 2.3.2. Control Intervention

The control intervention entailed watching an engaging, but emotionally neutral documentary (“When we left earth-the NASA missions” by Discovery entertainments, 2008) for the same duration as the experimental intervention. This has been commonly used in previous mental fatigue studies (42, 46). The duration of the control intervention was based on a pilot study which showed that the participants completed the 1500 Stroop trials within about an hour.

### 2.4. Subjective Assessments

The mental fatigue visual analogue scale (M-VAS) was used to assess the subjective feeling of mental fatigue before and after each intervention. The subjective mental and effort demands of the experimental and control interventions were assessed after each intervention using the relevant sub-sections of the National Aeronautics and Space Administration (NASA) Task Load Index.

### 2.5. Objective Assessment of Mental Fatigue

To assess objective mental fatigue (i.e., time related decrease in Stroop performance), the 1500 trials were divided into three blocks, each containing 500 trials during analysis. Measures of cognitive performance and executive functioning including the overall processing speed (overall reaction time-RT), and conflict cost were compared across the blocks. Conflict cost refers to the difference in reaction time, number of errors and number of missed trials between incongruent (high conflict) and congruent (low conflict) Stroop trials (47). Higher conflict cost is an indicator of lower executive function (47), and we expected the conflict cost to increase across the blocks as the participants become increasingly mentally fatigued. Similarly, the overall processing speed is expected to decrease (i.e., increase in overall reaction time) across the blocks as the participants become mentally fatigued.

### 2.6. Assessment of CSE and CCE

CSE and CCE were evaluated by measuring MEPs induced from M1 using single-pulse and paired-pulse TMS, respectively (48). This was achieved through the following procedure.

#### 2.6.1. Participants Positioning and Electromyography

The participants were seated comfortably on an adjustable chair, with their head well supported on the head rest, and the right arm relaxed in a pronated position on the arm rest of the chair (48). The first dorsal interosseous (FDI) was the muscle chosen for recording the MEP through surface electromyography (EMG) based on the strong corticospinal projection to spinal motor neuron pool innervating this muscle, and therefore the ease of stimulation and quality of the EMG signals (34). The location of the FDI was determined through palpation during a manually resisted abduction of the second digit by the participants (41). After determining the FDI location, the skin over the muscle was prepared by gently abrading the skin using medical grade sandpaper and then cleaning it using alcohol wipes, before placing the EMG electrodes. This was to ensure good surface contact and minimize skin resistance (41). The skin over the dorsum of the right hand was prepared in the same way. Two pre-gelled self-adhesive bipolar Ag/AgCl disposable surface electrodes were then placed over the FDI with an inter-electrode distance of 2 cm and a ground electrode was placed over the dorsum of the hand. EMG signals were filtered (10–500 Hz), amplified (× 1000) and sampled at 1000 Hz. All data were recorded on PC using LabChartTM software (ADInstruments, Australia) via a laboratory analogue-digital interface (PowerLab, ADInstruments, Australia) for later off-line analysis.

#### 2.6.2. Identifying hotspot and determination of the resting motor threshold

To locate the hotspot for the stimulation of the right FDI using TMS, the vertex (i.e., the Cz in the 10-20 electrode system) on the participants head was first located by measuring the mid-point between the nasion and the inion and the two preauricular areas using a soft tape measure (49). This was indicated with a non-permanent marker, followed by measuring 5cm laterally to the left of this marked point (49). This point was also marked, and the centre of TMS coil was then placed directly over the scalp on this area with its horizontal plane parallel to the participant’s head. A 70 mm figure of eight magnetic coil (MagVenture, Denmark), connected to a MagPro R30 stimulator (MagVenture, Denmark) was used for this study. The centre of the figure-of-eight coil was placed over the target area for effective stimulation, while its handle was aligned at approximately 45 degrees to the parasagittal plane and pointing backward (49, 50). Stimulations were then applied to elicit MEP with peak-to-peak amplitude of between 0.5-1millivolts (mV) in the target muscle (right FDI). The coil was carefully moved in each one of these four directions (i.e., anteriorly, posteriorly, medially, and laterally) eliciting 3 MEP in each site and taking note of the size of the MEP. The site that consistently produced largest MEPs was noted, marked, and used as the hotspot for the stimulation (49). The RMT, defined as the lowest TMS intensity that elicited MEP with a peak-to-peak amplitude of at least 50 microvolts in the resting FDI was then determined via stimulations applied at the hotspot (49). This was achieved via the parameter estimation by sequential testing (PEST) technique, which involves a stepwise suggestion of different intensities of stimulation by a software-motor threshold assessment tool (MTAT), until the RMT is finally determined (49). The TMS intensity was defined as a percentage of maximum stimulator output (%MSO) (49, 51). The 120% of the RMT was used as the intensity of stimulation to elicit the MEP for the assessment of CSE using single pulse TMS (48, 51).

To obtain the baseline mean MEP within the range of 1 mV ± 20% for the calculation of the SICI and ICF during paired pulse stimulation, another test intensity was determined by increasing the RMT until an intensity that produces MEP with peak-to-peak amplitude around 1 mV (51). This was called the 1mV test intensity.

#### 2.6.3. Single-Pulse TMS for assessment of CSE

Twenty-five single pulse stimuli at an inter-pulse interval of 6 seconds were applied over the hotspot, at an intensity of 120% of the RMT to elicit 25 consecutive MEPs in the resting FDI. The mean peak-to-peak amplitude of these MEP was used as the measure of CSE and was assessed before and immediately after both the experimental and the control interventions. Additionally, another 25 single pulses were applied over the hotspot using the 1mV test intensity earlier determined. The average peak-to-peak amplitude of the MEP elicited from this stimulation was used as the baseline for calculating the SICI and ICF (51).

#### 2.6.4. Paired pulse TMS for assessment of CCE

During the paired pulse stimulation, a conditioning stimulus was followed by a test stimulus after different time intervals to determine the SICI, LICI and ICF, which are the indices for assessment of CCE (48). For SICI and ICF, a subthreshold conditioning stimulus applied at 80% of the RMT was followed by a suprathreshold test stimulus at 1mV test intensity within an interval of 3 milliseconds (ms) (SICI) and 10ms (ICF) in a random order (51). Twenty-five MEPs were elicited each for SICI and ICF and the average peak-to-peak amplitude of each was calculated. This was then divided by the average peak-to-peak amplitude of the 25 MEPs earlier elicited via single-pulse stimulation using the 1mV test intensity, and then multiplied by 100 to obtain the percentage of inhibition in SICI and facilitation for ICF (51).

For LICI, a suprathreshold stimulus at 120% of the RMT was followed by a similar suprathreshold stimulus within an interval of 150 ms (51). Twenty-five MEPs were elicited using this technique and the average value of their peak-to-peak amplitude was then divided by the average of the peak-to-peak amplitude of the 25 MEP earlier elicited via single pulse stimulation at an intensity of 120% of the RMT. This value was then multiplied by 100 to obtain the LICI parameter (51).

### 2.7. Post Intervention Assessment of CSE and CCE

After both the experimental and the control interventions, the same procedure was followed to assess the CSE and CCE using single-pulse and paired-pulse TMS respectively. However, a new RMT and 1mV test intensity were determined (adjusted intensities) and used for the stimulation (51).

## 3. Statistical analysis

Statistical analyses were conducted using the Statistical Package for the Social Sciences (SPSS) version 28 (IBM Corp., Armonk, NY, USA). The data were first screened for normal distribution using the Shapiro–Wilk test and visual inspection of histograms.

Two-way (2 × 2) repeated measure analysis of variance (RM-ANOVA) was used to test the effect of intervention (mental fatigue and control) and time (pre and post) on the subjective feeling of mental fatigue (M-VAS score). Four similar two-way (2 × 2) RM-ANOVA were conducted to test the effect of intervention (mental fatigue and control) and time (pre and post) on the measures of CSE and CCE (SICI, ICF & LICI).

The measures of CSE and CCE were normalized to the pre value by dividing each value (pre and post values) with the pre value before statistical analyses (52). The normalized values are described by the symbol “*n*” written before the respective measures (*n*MEP, *n*SICI, *n*ICF and *n*LICI). A *n*MEP value ˃ 1 post intervention indicates increased CSE relative to before the intervention, and vice-versa. In the same way, a *n*ICF value ˃ 1 after the intervention indicate increased cortical facilitation compared to before the intervention, while values < 1 indicate the opposite. *n*SICI and *n*LICI values of < 1 and ˃ 1 after compared to before the intervention indicates an increase in cortical inhibition and a release of cortical inhibition respectively (52).

The normalization procedure was implemented to eliminate potential confounding factors in the statistical analyses that could stem from the inherent interindividual variability in the pretest values of the measures of cortical and corticospinal excitability (52). Analysing normalized values helps to mitigate any bias introduced by data from individual participants with notably low or high pretest values (52).

To examine objective mental fatigue due to the performance of the Stroop task, One-way RM-ANOVA was applied to the mean overall reaction time and the conflict cost (reaction time) data. The conflict cost data concerning number of missed trials and number of errors was not normally distributed. Therefore, a nonparametric Freidman test was used to examine objective mental fatigue in this data. Before interpreting RM ANOVA statistical outcomes, sphericity was verified by Mauchly’s test. When the assumption of sphericity was violated, Greenhouse-Geiser corrected significance values were used. Where main effects were found in RM-ANOVA analyses, they were further investigated using post-hoc pairwise comparisons with Bonferroni correction. Decline in cognitive performance over time (objective mental fatigue) is a necessary condition for both conceptual and operational definition of mental fatigue (13).

Therefore, it is critical to confirm that cognitive performance declined over time before making inference about the neural substrate underlining mental fatigue effects (13). However, objective mental fatigue may be subject to interindividual differences (13). In view of this, where the analysis of the mean Stroop task data of the participants above did not reveal objective mental fatigue, individual participants data was analyzed. This may reveal a fatigued subgroup that will enable proper investigation of the neural substrate underlining mental fatigue effects (13).

Finally, participants perceived workload (mental demand and effort subscales of the NASA-TLX Index) after the experimental and control interventions were compared using paired sample t-test. Significance was set at 0.05 for all the analyses.

## 4. Results

### 4.1. Subjective mental fatigue

There was no significant interaction between the type of intervention and time in the M-VAS score of the participants (F (1,14) = 3.197, p = 0.095, ηp2= 0.186) (Figure 3). Similarly, there was no main effect of intervention on the participant’s M-VAS score (F (1,14) = 1.847, p = 0.196, ηp2= 0.117). However, there was a main effect of time on the M-VAS score (F (1,14) = 24.925, p ˂ 0.001*, ηp2= 0.640). Post hoc comparison revealed that this was because the average M-VAS score increased from pre to post across both the mental fatigue and the control interventions, indicating increased subjective mental fatigue regardless of the type of intervention.

**Figure 3a-c.**
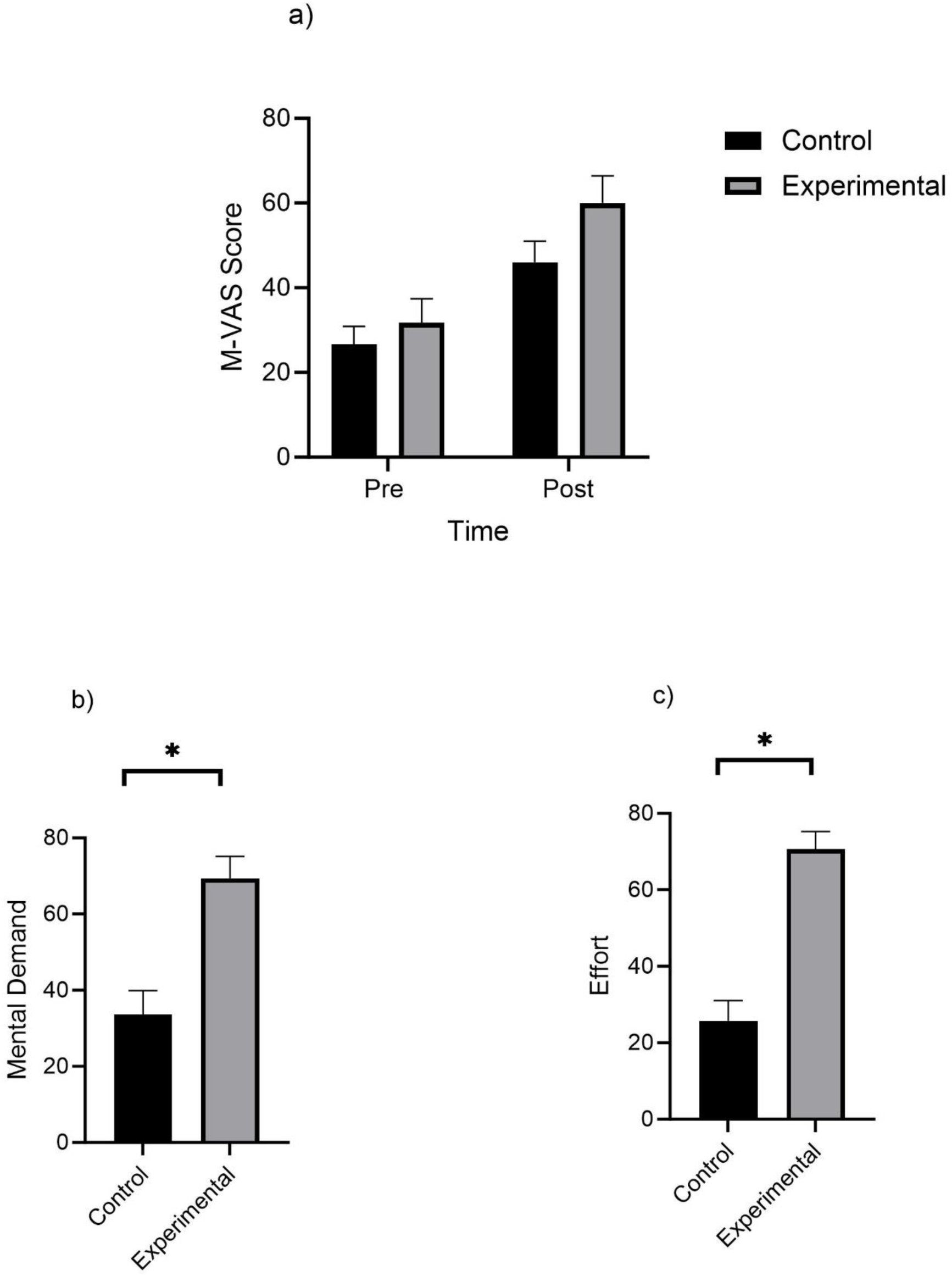
Results of the analyses showing the comparison between the mental fatigue and control conditions in a) subjective mental fatigue, b) perceived mental demand and c) perceived effort (* indicate a significant difference between the experimental and control conditions). Data is presented as mean and SEM.

### 4.2. Perceived workload

The participants perceived the performance of the Stroop task to be more effortful and mentally demanding than watching the documentary (p<0.05) (Figure 3).

### 4.3. Objective mental fatigue

We encountered technical computer error after the Stroop task performance by three participants which resulted in the loss of their Stroop task results. Consequently, only the Stroop task results of the 12 remaining participants were analysed. There were no significant main effect of block on any of the measures of Stroop task performance examined (Figure 4): Overall reaction time (F (1.342, 14.758) = 2.693, p = 0.115, ηp2= 0.197); conflict cost-reaction time (F (2, 22) = 0.876, p = 0.431, ηp2= 0.074); conflict cost-number of errors (χ2 (2) =1.319, p=0.517); and conflict cost-number of missed trials (χ2 (2) =2.098, p=0.350). This indicates that there was no evidence of objective mental fatigue due to the performance of the Stroop task in the participants.

**Figure 4a-d.**
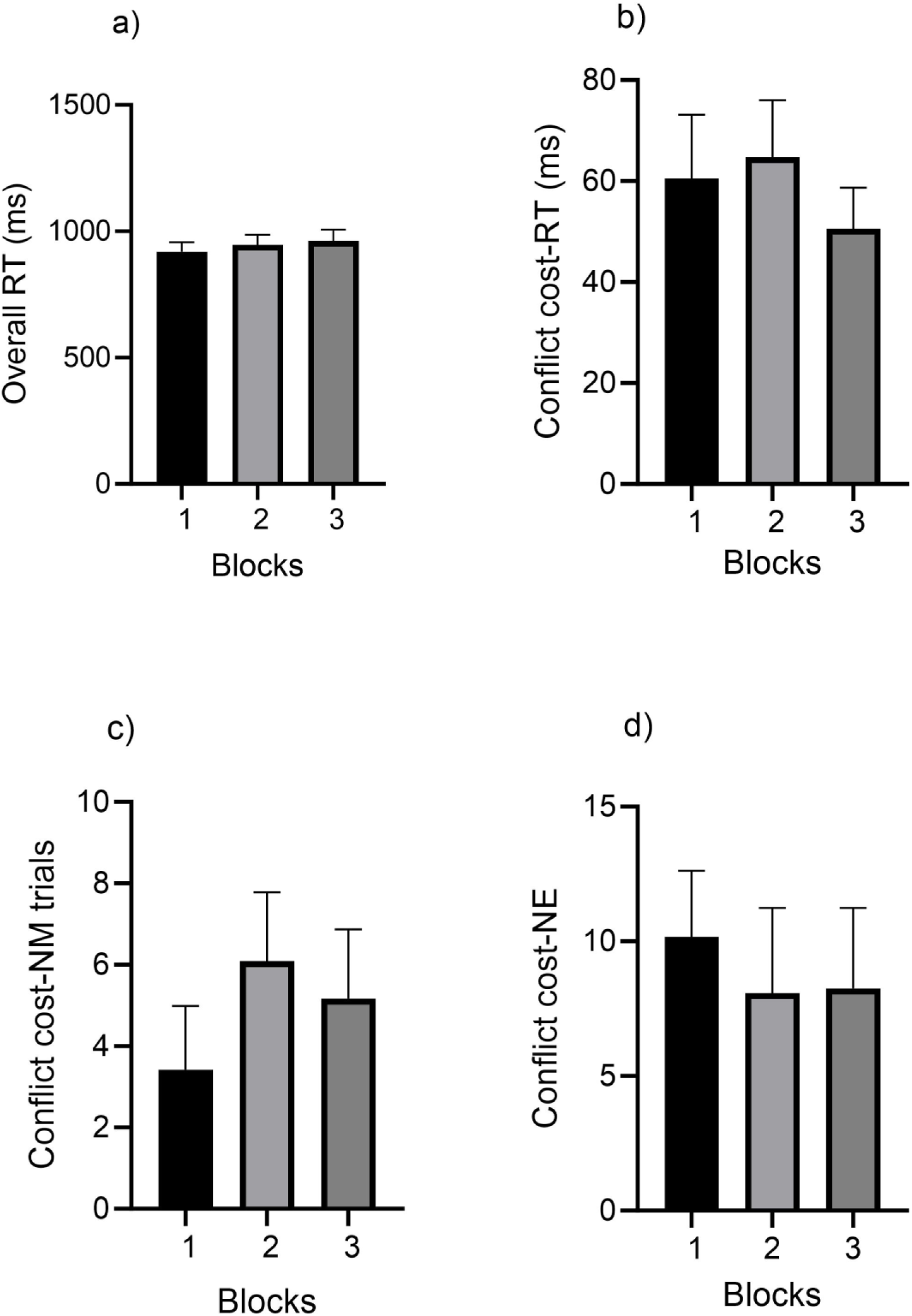
Time related change in measures of Stroop task performance; a) Overall reaction time b) Conflict cost reaction time c) Conflict cost number of missed trials and d) Conflict cost number of errors (objective mental fatigue assessment). Ms = milliseconds, NM = Number of missed trials, NE = Number of errors. Data is presented as mean and SEM.

### 4.4. CSE and CCE

Results of the analysis of the CSE and different indices of CCE (ICF, SICI and LICI) revealed no significant interaction between the type of intervention and time for all the measures (p˃0.05) (Table 1, Figure 5). Similarly, there was no significant main effect of intervention (p˃0.05) on any of the measures. However, there was a significant main effect of time on ICF (F (1,14) = 6.064, p = 0.027*, ηp2= 0.302) and LICI (F (1,14) = 5.233, p = 0.038*, ηp2= 0.272) (Table 1). Post hoc testing indicates that this was because averagely across both the mental fatigue and control interventions, the normalized ICF and LICI values were higher in the post compared to pre intervention periods (p<0.05).

**Figure 5a-d.**
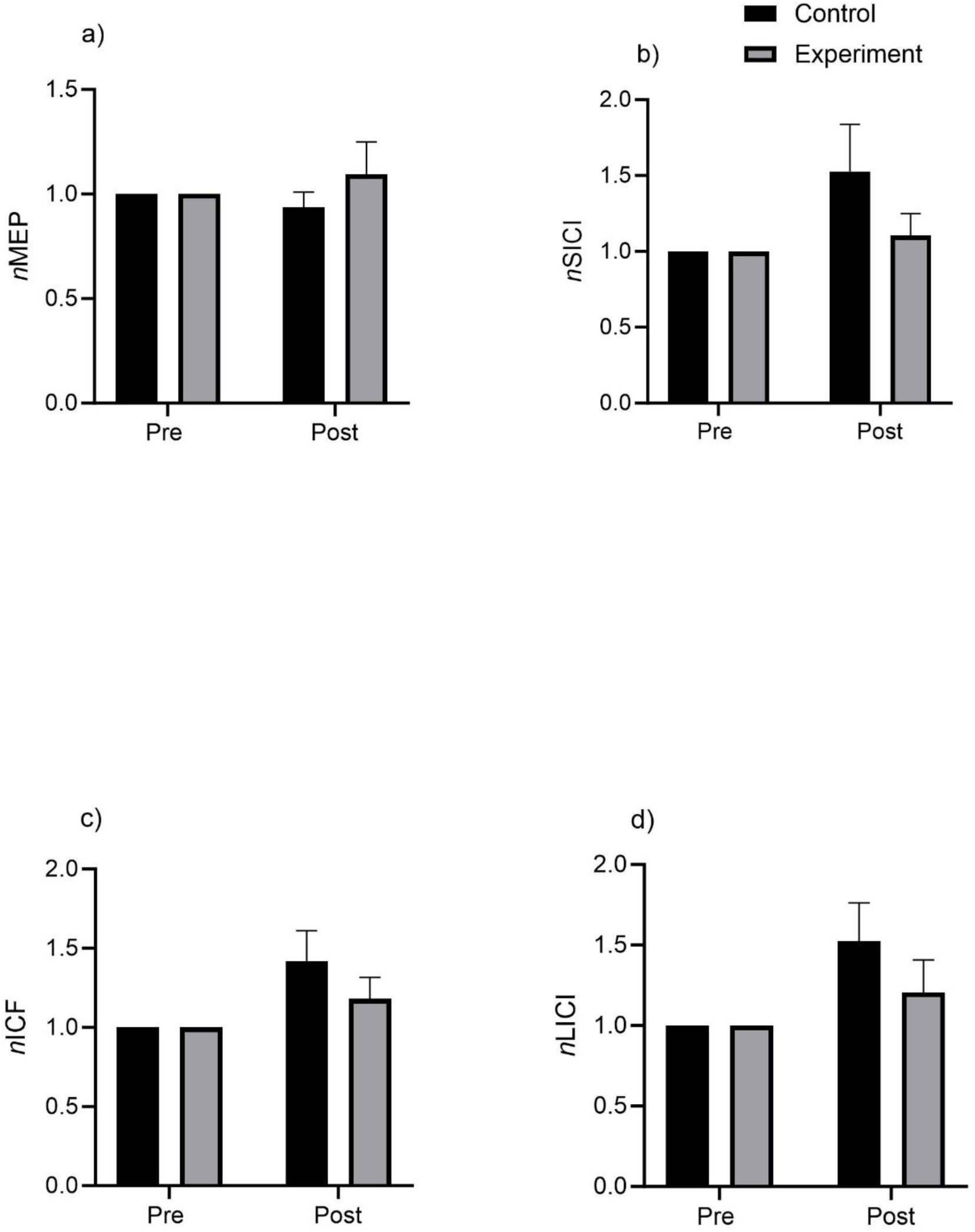
Normalized values for the measures corticospinal excitability (a) and cortico-cortical excitability (c-d) before and after the experimental and control conditions. MEP = Motor evoked potential, SICI = Short-interval intracortical inhibition, ICF = Intracortical facilitation, LICI = Long-interval intracortical inhibition. Data is presented as mean and SEM.

**Table 1.** Statistics-CSE and CCE outcomes.

| Outcomes | MAIN AND INTERACTION EFFECTS |  |  |
| --- | --- | --- | --- |
|  | Two-way interactions | Main effects |  |
|  | Exp-cond x time | Exp-cond | time |
| CSE | (F (1,14) = 0.698, p = 0.418, $\eta_p^2$= 0.047 | (F (1,14) = 0.698, p = 0.418, $\eta_p^2$= 0.047 | (F (1,14) = 0.038, p = 0.849, $\eta_p^2$= 0.003 |
| ICF | (F (1,14) = 1.122, p = 0.307, $\eta_p^2$= 0.074 | (F (1,14) = 1.122, p = 0.307, $\eta_p^2$= 0.074 | (F (1,14) = 6.064, p = 0.027*, $\eta_p^2$= 0.302 |
| SICI | (F (1,14) = 1.401, p = 0.256, $\eta_p^2$= 0.091 | (F (1,14) = 1.401, p = 0.256, $\eta_p^2$= 0.091 | (F (1,14) = 1.122, p = 0.307, $\eta_p^2$= 0.074 |
| LICI | (F (1,14) = 1.125, p = 0.307, $\eta_p^2$= 0.074 | (F (1,14) = 1.125, p = 0.307, $\eta_p^2$= 0.074 | (F (1,14) = 5.233, p = 0.038*, $\eta_p^2$= 0.272 |

### 4.5. Analysis of Stroop task performance changes overtime in individual participants

A previous study has shown that participants may respond differently to mental fatigue tasks (Stroop task in this study), whereby some may show decreased cognitive performance with time (objective mental fatigue) while others may not (13). The results above showed no evidence of objective mental fatigue when the whole participant set data was analysed. Similarly, there was no significant difference in the measures of CSE and CCE between the mental fatigue and the control interventions. In view of this, the overall processing speed (overall reaction time) of each participant was divided into three blocks and analysed using nonparametric Friedman test because the data was not normally distributed. The participants that showed decreased processing speed (increased overall RT) across the blocks were classified as a fatigued subgroup, while those that showed stable or increased processing speed (decreased RT) were classified as a non-fatigued subgroup, as done in a previously published study (13). One-way RM-ANOVA (data were normally distributed) were then applied to the mean overall processing speed and the conflict cost data in each subgroup to investigate objective mental fatigue. The effect size for the difference in measures of CSE and CCE between pre and post Stroop task performance was also calculated in each sub-group. This was done to explore how the performance of the Stroop task may affect these measures in the two sub-groups.

### 4.6. Results of individual participants data analysis

Results of the analysis of the change in the overall processing speed of the individual participants with time was summarized in Table 2. Six participants showed decreased processing speed (increased overall RT) with time, indicating objective mental fatigue. These are termed the fatigued sub-group. The remaining six participants showed either no significant change or an increase in processing speed (decreased overall RT). These are termed the non-fatigued subgroup.

**Table 2.** Results of the analysis for the changes in overall processing speed in individual participants across the blocks.

| Participants | Friedman test results | Multiple comparisons | P-value | Effect direction | Comment |
| --- | --- | --- | --- | --- | --- |
| Participant 4 | $\chi^2 (2) = 25.80$,<br>$p < 0.0001$ | Block 1 vs Block 2 | 0.001 | ↑ RT | Fatigued |
| | | Block 1 vs Block 3 | $< 0.0001$ | ↑ RT | |
| Participant 5 | $\chi^2 (2) = 28.94$,<br>$p < 0.0001$ | Block 1 vs Block 2 | 0.0003 | ↑ RT | Fatigued |
| | | Block 1 vs Block 3 | $< 0.0001$ | ↑ RT | |
| Participant 12 | $\chi^2 (2) = 40.03$,<br>$p < 0.0001$ | Block 1 vs Block 2 | $< 0.0001$ | ↑ RT | Fatigued |
|  |  | Block 1 vs Block 3 | 0.015 | ↑ RT |  |
| Participant 13 | $\chi^2 (2) = 15.38$, $p = 0.0005$ | Block 1 vs Block 2 | 0.1498 | ↑ RT | Fatigued |
|  |  | Block 1 vs Block 3 | 0.0003 | ↑ RT |  |
| Participant 14 | $\chi^2 (2) = 45.47$,<br>$p < 0.0001$ | Block 1 vs Block 2 | $< 0.0001$ | ↑ RT | Fatigued |
| | | Block 1 vs Block 3 | $< 0.0001$ | ↑ RT | |
| Participant 15 | $\chi^2 (2) = 21.39$,<br>$p < 0.0001$ | Block 1 vs Block 2 | 0.0008 | ↑ RT | Fatigued |
| | | Block 1 vs Block 3 | $< 0.0001$ | ↑ RT | |
| Participant 1 | $\chi^2 (2) = 11.53$, $p = 0.003$ | Block 1 vs Block 2 | 0.003 | ↓ RT | Non-Fatigued |
|  |  | Block 1 vs Block 3 | 0.04 | ↓ RT |  |
| Participant 2 | $\chi^2 (2) = 11.56$, $p = 0.003$ | Block 1 vs Block 2 | 0.0029 | ↓ RT | Non-Fatigued |
|  |  | Block 1 vs Block 3 | 0.9 | ↓ RT |  |
| Participant 3 | $\chi^2 (2) = 0.02104$, $p = 0.9895$ | NA | | ↓ RT | Non-Fatigued |
|  |  | NA |  | ↓ RT |  |
| Participant 6 | $\chi^2(2) = 12.49, p = 0.0019$ | Block 1 vs Block 2 | 0.002 | ↓ RT | Non-Fatigued |
|  |  | Block 1 vs Block 3 | 0.99 | ↓ RT |  |
| Participant 8 | $\chi^2(2) = 6.672, p = 0.035$ | Block 1 vs Block 2 | 0.99 | ↓ RT | Non-Fatigued |
|  |  | Block 1 vs Block 3 | 0.04 | ↓ RT |  |
| Participant 10 | $\chi^2(2) = 10.66, p = 0.0048$ | Block 1 vs Block 2 | 0.006 | ↓ RT | Non-Fatigued |
|  |  | Block 1 vs Block 3 | 0.04 | ↓ RT |  |

RT = Reaction time
NA = Not applicable
↓ = Decreased
↑ = Increased

Analysis of the mean overall processing speed separately in the two subgroups showed that, while the mean overall processing speed decreases (i.e. the overall reaction time increases) with time in the fatigued subgroup (F (2, 10) = 23.867, p < 0.001, ηp2= 0.827) (Figure 6), the non-fatigued subgroup showed an opposite pattern (increased processing speed from block 1 to 2, and no significant difference between block 3 and 1) (F (2, 10) = 9.101, p = 0.006, ηp2= 0.645). Analysis of conflict cost parameters did not show significant change with time in both subgroups. However, descriptively, the fatigued subgroup showed deterioration of the conflict cost measures with time (i.e., increased conflict cost), while the non-fatigue group showed improvement (Figure 7).

**Figure 6.**
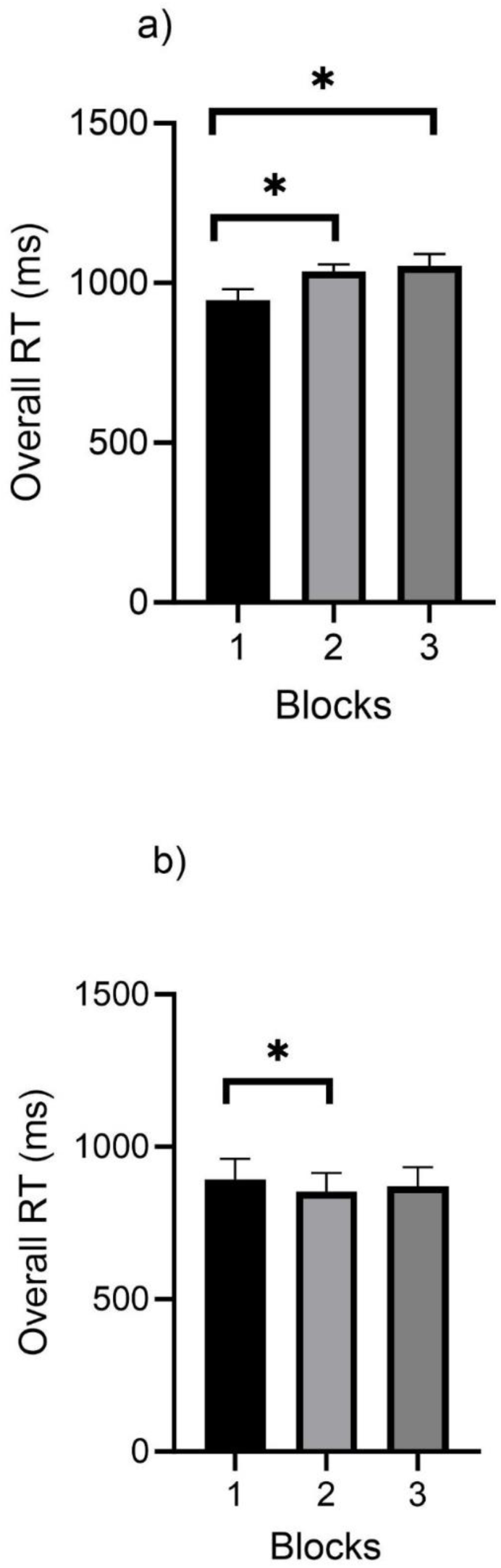
Results of the analyses showing the changes in the overall reaction time with time (objective mental fatigue assessment) in a) the fatigued subgroup and b) the non-fatigued subgroup. RT = Reaction time. Data is presented as mean and SEM.

**Figure 7a&b.**
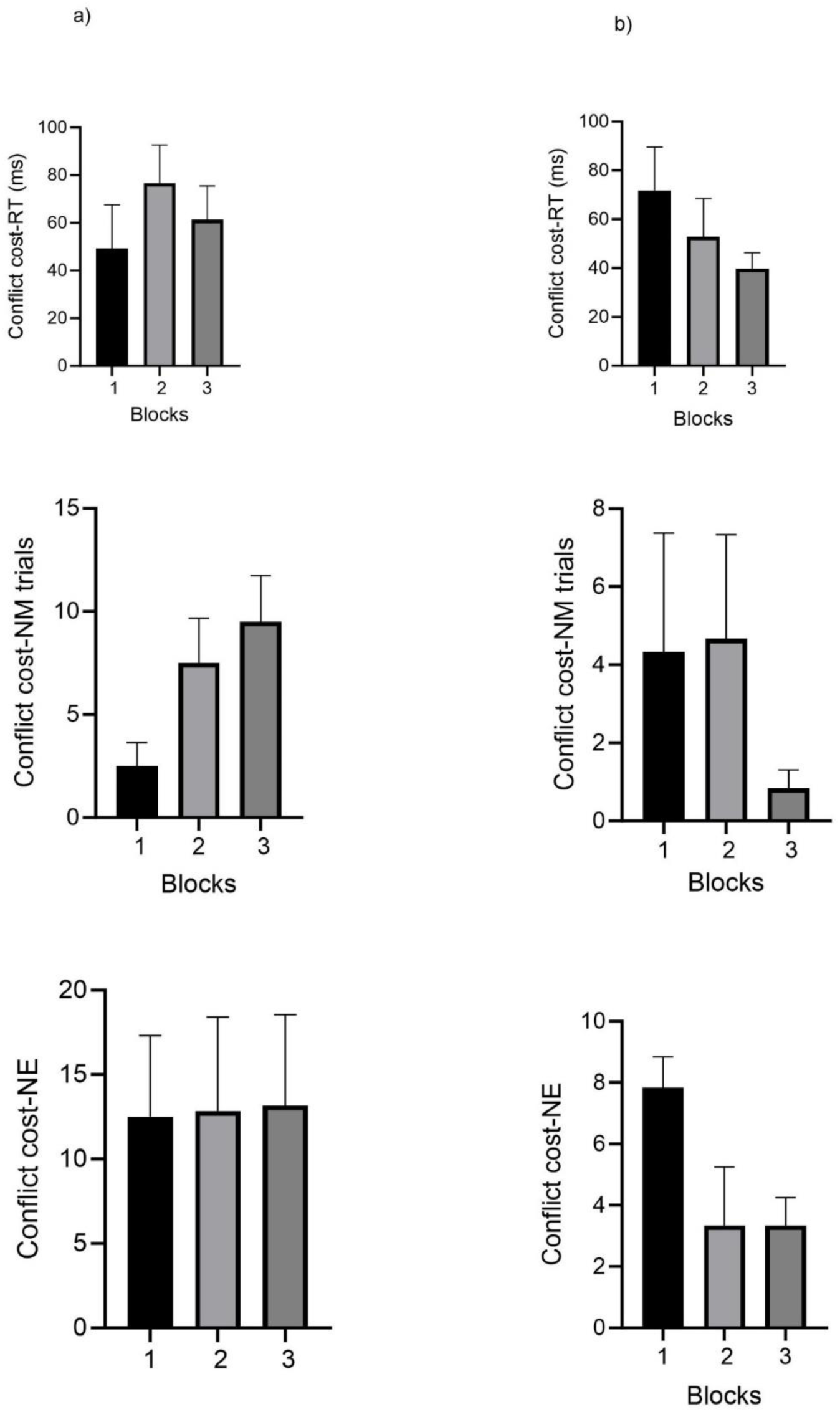
Results of the analyses showing the changes in measures of conflict cost with time (objective mental fatigue assessment) in a) the fatigued subgroup and b) the non-fatigued subgroup. NM = Number of missed trials, NE = Number of errors, RT = Reaction time. Data is presented as mean and SEM.

Effect sizes and 95% confidence level (CL) calculated for the difference in measures of CSE and CCE from before to after the Stroop task also showed different effects direction on some of the measures in the two sub-groups. Although not statistically significant, but descriptively, the ICF and SICI parameters decreased from pre to post in the fatigued subgroup (ICF: d = −0.73, 95% CI −1.90 to 0.44; SICI: d = −0.37, 95% CI −1.51 to 0.78), while they increased in the non-fatigued subgroup (ICF: d = 0.25, 95% CI −0.89 to 1.39; SICI: d = 0.05, 95% CI −1.08 to 1.18) (Figure 8a&b).

**Fig 8a.**
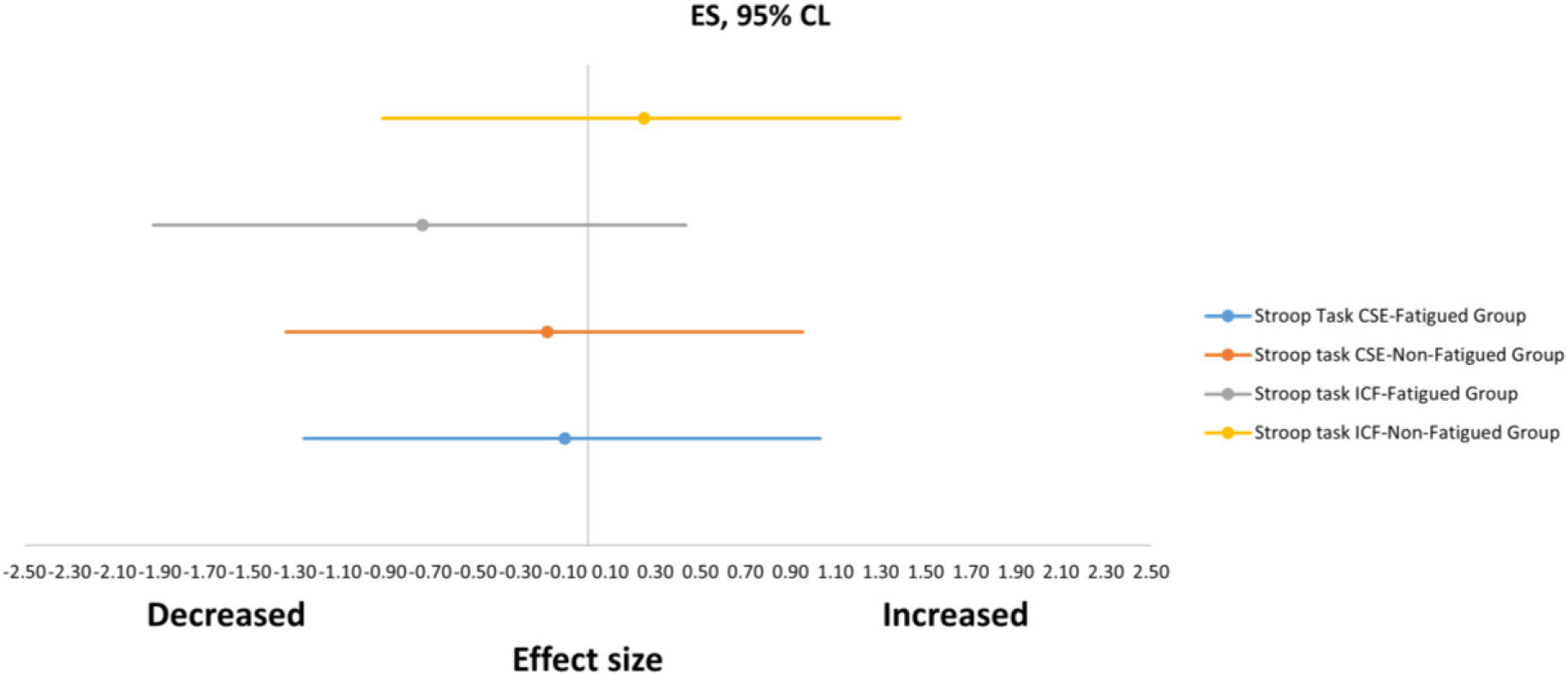
Effect size calculated for the difference in corticospinal excitability (CSE) and ICF before and after Stroop task performance in the fatigued and the non-fatigued subgroups.

**Fig 8b.**
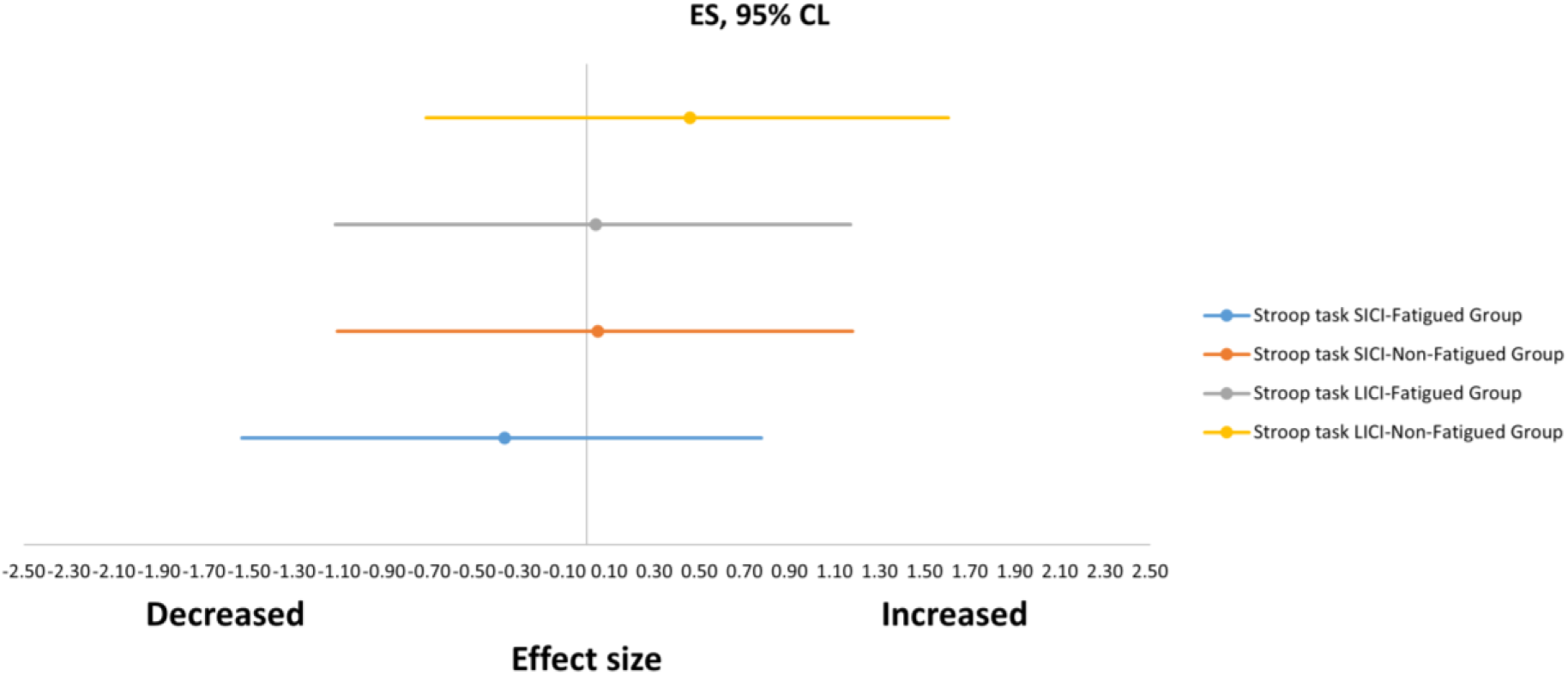
Effect size calculated for the difference in SICI and LICI before and after Stroop task performance in the fatigued and the non-fatigued subgroups.

## 5. Discussion

The current study was conducted to investigate the effects of mental fatigue induced by prolonged cognitive performance on CSE and CCE. This was achieved by comparing measures of CSE (peak-to-peak amplitude of single pulse TMS-Induced MEP) and CCE (ICF, SICI and LICI) before and after a prolonged cognitive task (Stroop task) and a control task (watching a documentary) in a group of healthy adults. Any change in the measures of CSE and CCE will depend on successful induction of mental fatigue, especially objective mental fatigue in the participants (13). It is therefore crucial to first discuss the findings of the current study regarding whether mental fatigue was successfully induced in the participants or not, before the discussion about the potential effects on CSE and CCE.

### 5.1. Subjective and Objective Mental Fatigue

In the mental fatigue literature, both subjective and objective measures are commonly used to examine the successful induction of mental fatigue (42, 46, 53). Using a mental fatigue visual analogue scale (M-VAS) to assess subjective feeling of mental fatigue, the result of the current study showed no significant difference in the M-VAS score after the two interventions. However, there was a significant main effect of time, indicating an increased feeling of mental fatigue from pre to post regardless of the type of the intervention. Therefore, subjective feeling of mental fatigue can be said to increase after both the Stroop task performance and watching the documentary. The effort and mental demand sub-scales of the NASA-TLX index used to examine the perceived mental demand and effort associated with the performance of the two interventions have shown a different result. The participants perceived the performance of the Stroop task to be more effortful and mentally demanding than watching the documentary. This indicates that, although the two tasks differed in the perceived workload associated with their performance, both resulted in subjective feelings of mental fatigue. This finding is in line with the results of a recent study which suggests that watching a documentary may not constitute a suitable control in mental fatigue studies, as it also results in subjective feeling of mental fatigue in the participants (54). Interestingly, the study has shown that watching a documentary results in feeling of mental fatigue by causing under-arousal and sleepiness (54). Therefore, it is plausible that the two tasks differed in their perceived workload, but both resulted in subjective mental fatigue, albeit through different mechanisms. Perhaps, the performance of the Stroop task resulted in subjective mental fatigue due to overload, while watching the documentary caused subjective mental fatigue due to boredom and underload (53–55). The implication of this finding is that, future studies should consider using a more neutral control when investigating the effects of mental fatigue on CSE and CCE (54).

Regarding the objective mental fatigue assessed by examining the time related change in the performance of the Stroop task, there was no significant deterioration in any of the indices of Stroop task performance with time when the mean data of all the participants was analysed. This suggests that there was no evidence of objective mental fatigue in the participants. This is in accordance with the findings of similar previous studies that used the Stroop task containing predominantly incongruent trials to induce mental fatigue (42, 46). Compared to simpler cognitive tasks, the performance of more difficult types of cognitive tasks requiring executive control, such as incongruent Stroop trials have been shown to remain stable with time in several previous studies (42, 46, 53, 55–57). Perhaps, the difficulty of the tasks aroused participants to apply adequate effort and concentration on the tasks to maintain performance (53). Alternatively, the familiarization trials carried out before the tasks were not enough to ensure adequate learning due to the difficulty of the tasks (39). Therefore, ongoing learning during the prolonged performance of the tasks may counteract the expected fatigue related deterioration of performance, resulting in overall stable performance (39, 55). In addition to the above mentioned factors, interindividual differences in susceptibility to objective mental fatigue may also play a role (13). The data from a group of participants that are less susceptible to objective mental fatigue during prolonged performance of cognitively demanding task may counteract that of the more susceptible group, thereby masking the overall objective mental fatigue (13). Indeed, when individual participants overall processing speed was examined, a subgroup that showed a deterioration of processing speed or objective mental fatigue with time emerged. Although not statistically significant, the evolution of conflict cost with time was also descriptively in accordance with development of objective mental fatigue in this subgroup. This is in line with the findings of a previous study that applied the same approach (13). Interestingly, the type of task used in that study also required executive function (conflict resolution) similar to the majority of the incongruent Stroop trials used in the current study (13). Therefore, interindividual variability in the susceptibility to mental fatigue should be considered during objective mental fatigue analysis to separate the participants that are objectively mentally fatigued from those that are not (13).

Overall, evidence of subjective mental fatigue appeared in the participants of the current study after both the experimental and control interventions. In terms of objective mental fatigue during the Stroop task performance, a subgroup of the participants was mentally fatigued, while the remainder were not.

### 5.2. Effect on CSE and CCE

The results of the current study regarding the effects of mental fatigue on M1 excitability did not show significant differences in the measures of CSE (single pulse MEP peak-to-peak amplitude) and CCE (ICF, SICI and LICI) between the mental fatigue and the control interventions when the mean data of all the participants were analysed. This is not surprising, considering the lack of difference in subjective mental fatigue between the two interventions, as well as the absence of objective mental fatigue following the Stroop task performance. In a similar previous study by Kowalski and associates where the participants carried out a psychomotor vigilance task (PVT) and watched documentary as the mental fatigue and control interventions respectively, no significant effect of time or condition was found on MEP peak-to-peak amplitude assessed using single pulse TMS (28). However, cortical silent period, a measure of the activity of cortical inhibitory circuits mediated via Gamma-aminobutyric acid B (GABA_B_) receptors (34), was found to become longer after both carrying out the PVT and watching the documentary (28). The PVT performance in this study deteriorated with time, indicating objective mental fatigue (28). The findings from the study by Kowalski and colleagues have important relationship with the results of the current study. First, it has shown that, MEP amplitude assessed using single pulse MEP may not necessarily change due to mental fatigue, but other measures specifically related to cortical circuits may. Indeed, after a further analysis that separated the participants in the current study into objectively mentally fatigued and non-fatigued subgroups, measures of cortical excitability (ICF and SICI) decreased from pre to post Stroop task, albeit only descriptively in the fatigued subgroup alone. This suggests that prolonged cognitive performance leading to objective mental fatigue may result in increased cortical inhibition, without necessarily affecting the corticospinal excitability. Other studies that examined the changes in MEP amplitude (a measure representing corticospinal excitability) before and after prolonged cognitive tasks that objectively induced mental fatigue also did not find any significant change (30, 31). However, increase in the cortical silent period duration approached significance in one of these studies that examined cortical silent period duration in addition to MEP amplitude (31). This further supports the notion that mental fatigue is likely to affect cortical circuits, particularly causing increased inhibition, because a lengthened cortical silent period also reflects increased cortical inhibition. The MEP amplitude assessed using single pulse TMS (a measure of corticospinal excitability) depends on the excitability of the cortical neurons, the spinal motor neurons, the muscle as well as the integrity of the corticospinal tract (34). Being a phenomenon arising primarily from the brain (38, 39), mental fatigue may only directly affect the CCE, without necessarily causing direct effect on spinal motor neurons or the muscles. This may be the reason why the effects of mental fatigue is more likely to be observed on measures of M1 excitability specifically related to the cortical circuits, not the single pulse TMS induced MEP peak-to-peak amplitude, which is a product of the excitability of both cortical and peripheral structures (34). However, this does not completely rule out potential changes in CSE due to mental fatigue, because the CSE partly depend on the CCE and changes in CCE could therefore cause the CSE to change too. Indeed, the current study and the previously mentioned studies have both induced mental fatigue through performance of cognitive tasks that are relatively shorter in duration (30 minutes to two hours) compared to other studies that used longer task duration (up to six hours) and shown clear changes in activity of cognitive control areas of the brain (38, 39). Perhaps, the use of cognitive tasks of similarly long duration may induce mental fatigue strong enough to cause significant and clear changes in CCE and subsequently the CSE.

This is an important question that should be investigated in future studies. In any case, the tendency of mental fatigue to bring about increased cortical inhibition is in line with the findings from the physical fatigue literature (58, 59). Research on the central mechanisms underlying physical fatigue has revealed that a crucial factor in the development of physical fatigue is the heightened cortical inhibition resulting in reduced CSE (58, 59).

The second point showing a similarity or relationship between the findings in the study by Kowalski and colleagues mentioned above and the current study, is the observation that the cortical silent period increased after both the mental fatigue and the control interventions (28). This supports our finding and that of another previous study that showed that prolonged watching of a documentary also results in mental fatigue (54). In fact, the study by Kowalski re-affirmed this by showing that mental fatigue resulting from watching a documentary produced increased cortical inhibition similar to that seen after a prolonged cognitive task (28). Notably, the cognitive task in the study by Kowalski and associates results in objective mental fatigue (28). Therefore, mentally induced fatigue may lead to increased cortical inhibition regardless of how it was induced. Perhaps, if objective mental fatigue were also observed due to the Stroop task when the entire participant dataset was analysed in the current study, similar increased cortical inhibition might occur regardless of the type of intervention. Overall, this suggests that a more neutral control condition needs to be developed to study the impact of mental fatigue due to prolonged cognitive performance on different variables including the M1 excitability (54). The current practice of asking participants to watch a documentary may bring about a confounding effect due to the mental fatigue induced by this process (54).

## 6. Limitations

The present study is subject to several limitations that warrant cautious interpretation of the findings. Primarily, the small overall sample size of 15 participants implies limited statistical power, potentially influencing the robustness of the analysis. Additionally, the supplementary analysis, which identified a subset of participants experiencing objective mental fatigue during the Stroop task, relied on even smaller sample size. Also, the findings relating to the increased intracortical inhibition due to mental fatigue were only descriptive and results from a pre-to-post Stroop comparison. This suggests that strong conclusions cannot be drawn from these findings. Finally, the participants in the current study are young and healthy. Different results may be observed when a population more vulnerable to mental fatigue such as older adults or individuals with neurological diseases were investigated.

## 7. Direction for future research

Exploring the impact of prolonged cognitive activity-induced mental fatigue on CSE or CCE represents a critical research area laying the groundwork for developing interventions that may mitigate the consequences of mental fatigue on physical performance. To enhance the robustness of these investigations, it is recommended that mental fatigue be induced in settings that mimic real-life conditions, utilizing tasks relevant to participants’ daily, occupational, or social activities. Conducting transcranial magnetic stimulation (TMS) assessments before and after a typical workday or social activity and implementing neutral control conditions can offer valuable insights into the nuanced relationship between mental fatigue and M1 excitability. Additionally, employing diverse subjective and objective measures to assess mental fatigue, acknowledging interindividual variability, and examining physical performance in various contexts are crucial for comprehensive analysis and interpretation. Lastly, expanding this research to clinical populations at higher risk of experiencing mental fatigue can further broaden the clinical applications of these findings.

## 8. Clinical implication

The trend towards increases in cortical inhibition observed due to mental fatigue implies that non-invasive brain stimulation techniques enhancing cortical excitability, such as anodal transcranial direct current stimulation, might prove effective in alleviating mental fatigue and its impact on physical performance across both healthy and clinical populations, assuming this finding is substantiated in subsequent research.

## 9. Conclusion

The study results revealed no difference in cortical and corticospinal excitability following either the mental fatigue (extended Stroop task engagement) or control (watching a documentary) intervention. This outcome is likely attributed to the confounding effects of mental fatigue induced by the control condition, along with the absence of objective mental fatigue, likely resulting from interindividual variability following the Stroop task performance. Further exploratory analysis, controlling for these factors, suggests the potential of mental fatigue to induce increased cortical inhibition in the M1, although this requires validation in future studies with larger sample sizes.

## Statements and Declarations

### Competing interests

All the authors certify that they have no affiliations with or involvement in any organization or entity with any financial interest or non-financial interest in the subject matter or materials discussed in this manuscript.

### Funding

This research did not receive any specific grant from funding agencies in the public, commercial, or not-for-profit sectors.

### Availability of data and materials

All data generated or analyzed during this study is available and can be provided if required.

### Consent to Participate

Not applicable.

### Consent for Publication

Not applicable.

### Authors contribution

Abubakar Tijjani Salihu: Conceptualization, Methodology, Formal Analysis, Data curation, writing-original draft preparation.

Shapour Jaberzadeh: Conceptualization, Methodology, writing-Review, and editing.

Keith D. Hill: Conceptualization, Methodology, writing-Review, and editing.

Maryam Zoghi: Conceptualization, Methodology, writing-Review, and editing.

## Notes

### Competing Interest Statement

The authors have declared no competing interest.

### Clinical Trial

Project ID: 27394

### Funding Statement

This study did not receive any funding

### Author Declarations

Etthics commitee of Monash University gave ethical approval for this work

## References

1. Boksem MA, Tops M. Mental fatigue: costs and benefits. Brain research reviews. 2008;59(1):125–39.

2. Caldwell JA, Caldwell JL, Thompson LA, Lieberman HR. Fatigue and its management in the workplace. Neuroscience & Biobehavioral Reviews. 2019;96:272–89.

3. Then FS, Luck T, Luppa M, Arélin K, Schroeter ML, Engel C, et al. Association between mental demands at work and cognitive functioning in the general population–results of the health study of the Leipzig research center for civilization diseases (LIFE). Journal of occupational medicine and toxicology. 2014;9(1):1–13.

4. Ahmed S, Rasul ME. Examining the association between social media fatigue, cognitive ability, narcissism and misinformation sharing: cross-national evidence from eight countries. Scientific Reports. 2023;13(1):15416.

5. Fortes LS, Gantois P, de Lima-Junior D, Barbosa BT, Ferreira MEC, Nakamura FY, et al. Playing videogames or using social media applications on smartphones causes mental fatigue and impairs decision-making performance in amateur boxers. Applied Neuropsychology: Adult. 2023;30(2):227–38.

6. Pignatiello GA, Martin RJ, Hickman Jr RL. Decision fatigue: A conceptual analysis. Journal of health psychology. 2020;25(1):123–35.

7. Russell S, Halson SL, Jenkins DG, Rynne SB, Roelands B, Kelly VG. Thinking About Elite Performance: The Experience and Impact of Mental Fatigue in Elite Sport Coaching. International Journal of Sports Physiology and Performance. 2023;1(aop):1–7.

8. Russell S, Jenkins D, Rynne S, Halson SL, Kelly V. What is mental fatigue in elite sport? Perceptions from athletes and staff. European journal of sport science. 2019;19(10):1367–76.

9. Maisel P, Baum E, Donner-Banzhoff N. Fatigue as the chief complaint: epidemiology, causes, diagnosis, and treatment. Deutsches Ärzteblatt International. 2021;118(33-34):566.

10. Wilson J, Morgan S, Magin P, van Driel M. Fatigue-a rational approach to investigation. Australian family physician. 2014;43(7):457–61.

11. Skarpsno ES, Nilsen TIL, Sand T, Hagen K, Mork PJ. Work-related mental fatigue, physical activity and risk of insomnia symptoms: Longitudinal data from the Norwegian HUNT Study. Behavioral Sleep Medicine. 2020;18(4):488–99.

12. Ricci JA, Chee E, Lorandeau AL, Berger J. Fatigue in the US workforce: prevalence and implications for lost productive work time. Journal of occupational and environmental medicine. 2007:1–10.

13. Holtzer R, Shuman M, Mahoney JR, Lipton R, Verghese J. Cognitive fatigue defined in the context of attention networks. Aging, Neuropsychology, and Cognition. 2010;18(1):108–28.

14. Marcora SM, Staiano W, Manning V. Mental fatigue impairs physical performance in humans. Journal of applied physiology. 2009.

15. Lou J-S. Physical and mental fatigue in Parkinson’s disease. Drugs & aging. 2009;26(3):195–208.

16. Chaudhuri A, Behan PO. Fatigue and basal ganglia. Journal of the neurological sciences. 2000;179(1-2):34–42.

17. Linnhoff S, Fiene M, Heinze H-J, Zaehle T. Cognitive fatigue in multiple sclerosis: an objective approach to diagnosis and treatment by transcranial electrical stimulation. Brain Sciences. 2019;9(5):100.

18. Hubacher M, Calabrese P, Bassetti C, Carota A, Stöcklin M, Penner I-K. Assessment of post-stroke fatigue: the fatigue scale for motor and cognitive functions. European neurology. 2012;67(6):377–84.

19. Mozuraityte K, Stanyte A, Fineberg NA, Serretti A, Gecaite-Stonciene J, Burkauskas J. Mental fatigue in individuals with psychiatric disorders: a scoping review. International Journal of Psychiatry in Clinical Practice. 2022:1–10.

20. Sadeghniiat-Haghighi K, Yazdi Z. Fatigue management in the workplace. Industrial psychiatry journal. 2015;24(1):12.

21. Brown DM, Graham JD, Innes KI, Harris S, Flemington A, Bray SR. Effects of prior cognitive exertion on physical performance: A systematic review and meta-analysis. Sports Medicine. 2020;50:497–529.

22. Habay J, Uylenbroeck R, Van Droogenbroeck R, De Wachter J, Proost M, Tassignon B, et al. Interindividual Variability in Mental Fatigue-Related Impairments in Endurance Performance: A Systematic Review and Multiple Meta-regression. Sports medicine-open. 2023;9(1):1–27.

23. Van Cutsem J, Marcora S, De Pauw K, Bailey S, Meeusen R, Roelands B. The effects of mental fatigue on physical performance: a systematic review. Sports medicine. 2017;47(8):1569–88.

24. Sun H, Soh KG, Roslan S, Wazir MRWN, Soh KL. Does mental fatigue affect skilled performance in athletes? A systematic review. PLoS One. 2021;16(10):e0258307.

25. Brahms M, Heinzel S, Rapp M, Mückstein M, Hortobágyi T, Stelzel C, et al. The acute effects of mental fatigue on balance performance in healthy young and older adults–A systematic review and meta-analysis. Acta psychologica. 2022;225:103540.

26. Grobe S, Kakar RS, Smith ML, Mehta R, Baghurst T, Boolani A. Impact of cognitive fatigue on gait and sway among older adults: A literature review. Preventive medicine reports. 2017;6:88–93.

27. Lew FL, Qu X. Effects of mental fatigue on biomechanics of slips. Ergonomics. 2014;57(12):1927–32.

28. Kowalski KL, Tierney BC, Christie AD. Mental fatigue does not substantially alter neuromuscular function in young, healthy males and females. Physiology & Behavior. 2022;253:113855.

29. Yip DW, Lui F. Physiology, motor cortical. 2019.

30. Terentjeviene A, Maciuleviciene E, Vadopalas K, Mickeviciene D, Karanauskiene D, Valanciene D, et al. Prefrontal cortex activity predicts mental fatigue in young and elderly men during a 2 h “Go/NoGo” task. Frontiers in neuroscience. 2018;12:620.

31. Morris AJ, Christie AD. The effect of mental fatigue on neuromuscular function is similar in young and older women. Brain sciences. 2020;10(4):191.

32. Derosière G, Billot M, Ward ET, Perrey S. Adaptations of motor neural structures’ activity to lapses in attention. Cerebral cortex. 2015;25(1):66–74.

33. Nakashima A, Moriuchi T, Matsuda D, Hasegawa T, Nakamura J, Anan K, et al. Corticospinal excitability during motor imagery is diminished by continuous repetition-induced fatigue. Neural Regeneration Research. 2021;16(6):1031–6.

34. Frazer A, Kidgell D. TMS-Induced Motor Evoked Potentials: Definitions and Physiology. A Closer Look at Motor-Evoked Potential. 2019:1–14.

35. Zewdie E, Kirton A. TMS basics: single and paired pulse neurophysiology. Pediatric Brain Stimulation: Elsevier; 2016. p. 3–22.

36. Zoghi M, Pearce SL, Nordstrom MA. Differential modulation of intracortical inhibition in human motor cortex during selective activation of an intrinsic hand muscle. The Journal of physiology. 2003;550(3):933–46.

37. Jaberzadeh S, Zoghi M. TMS Protocols for Assessment of Intracortical Facilitation. A Closer Look at Motor-Evoked Potential: Nova Science Publishers; 2019. p. 197–215.

38. Blain B, Hollard G, Pessiglione M. Neural mechanisms underlying the impact of daylong cognitive work on economic decisions. Proceedings of the National Academy of Sciences. 2016;113(25):6967–72.

39. Wiehler A, Branzoli F, Adanyeguh I, Mochel F, Pessiglione M. A neuro-metabolic account of why daylong cognitive work alters the control of economic decisions. Current Biology. 2022;32(16):3564–75. e5.

40. Veale JF. Edinburgh handedness inventory–short form: a revised version based on confirmatory factor analysis. Laterality: Asymmetries of Body, Brain and Cognition. 2014;19(2):164–77.

41. Pellegrini M, Zoghi M, Jaberzadeh S. The effect of transcranial magnetic stimulation test intensity on the amplitude, variability and reliability of motor evoked potentials. Brain Research. 2018;1700:190–8.

42. Verschueren J, Tassignon B, Proost M, Teugels A, Roelands B, Verhagen E, et al. Does mental fatigue negatively affect outcomes of functional performance tests? Medicine and science in sports and exercise. 2020;52(9):2002–10.

43. Pascoe AJ, Haque ZZ, Samandra R, Fehring DJ, Mansouri FA. Dissociable effects of music and white noise on conflict-induced behavioral adjustments. Frontiers in Neuroscience. 2022;16:858576.

44. Stoet G. PsyToolkit: A software package for programming psychological experiments using Linux. Behavior research methods. 2010;42:1096–104.

45. Stoet G. PsyToolkit: A novel web-based method for running online questionnaires and reaction-time experiments. Teaching of Psychology. 2017;44(1):24–31.

46. Tassignon B, Verschueren J, De Pauw K, Roelands B, Van Cutsem J, Verhagen E, et al. Mental fatigue impairs clinician-friendly balance test performance and brain activity. Translational Sports Medicine. 2020;3(6):616–25.

47. Mansouri FA, Tanaka K, Buckley MJ. Conflict-induced behavioural adjustment: a clue to the executive functions of the prefrontal cortex. Nature Reviews Neuroscience. 2009;10(2):141–52.

48. Pellegrini M, Zoghi M, Jaberzadeh S. Can genetic polymorphisms predict response variability to anodal transcranial direct current stimulation of the primary motor cortex? European Journal of Neuroscience. 2021;53(5):1569–91.

49. Dissanayaka T, Zoghi M, Farrell M, Egan G, Jaberzadeh S. Methods for Determination of Motor Threshold. Closer Look at Motor-Evoked Potential: Nova Science Publishers; 2019. p. 15–31.

50. Groppa S, Oliviero A, Eisen A, Quartarone A, Cohen L, Mall V, et al. A practical guide to diagnostic transcranial magnetic stimulation: report of an IFCN committee. Clinical Neurophysiology. 2012;123(5):858–82.

51. Behrangrad S, Zoghi M, Kidgell D, Mansouri F, Jaberzadeh S. The effects of concurrent bilateral anodal tDCS of primary motor cortex and cerebellum on corticospinal excitability: a randomized, double-blind sham-controlled study. Brain Structure and Function. 2022;227(7):2395–408.

52. Hinder MR, Schmidt MW, Garry MI, Carroll TJ, Summers JJ. Absence of cross-limb transfer of performance gains following ballistic motor practice in older adults. Journal of Applied Physiology. 2011;110(1):166–75.

53. Shigihara Y, Tanaka M, Ishii A, Tajima S, Kanai E, Funakura M, et al. Two different types of mental fatigue produce different styles of task performance. Neurology, Psychiatry and Brain Research. 2013;19(1):5–11.

54. O’Keeffe K, Hodder S, Lloyd A. A comparison of methods used for inducing mental fatigue in performance research: Individualised, dual-task and short duration cognitive tests are most effective. Ergonomics. 2020;63(1):1–12.

55. Salihu AT, Usman JS, Hill KD, Zoghi M, Jaberzadeh S. Mental fatigue does not affect static balance under both single and dual task conditions in young adults. Experimental brain research. 2023:1–16.

56. Moeller S, Tomasi D, Honorio J, Volkow N, Goldstein R. Dopaminergic involvement during mental fatigue in health and cocaine addiction. Translational psychiatry. 2012;2(10):e176–e.

57. Ackerman PL, Kanfer R. Test length and cognitive fatigue: an empirical examination of effects on performance and test-taker reactions. Journal of Experimental Psychology: Applied. 2009;15(2):163.

58. Weavil JC, Amann M. Corticospinal excitability during fatiguing whole body exercise. Progress in brain research. 2018;240:219–46.

59. Gruet M, Temesi J, Rupp T, Levy P, Millet G, Verges S. Stimulation of the motor cortex and corticospinal tract to assess human muscle fatigue. Neuroscience. 2013;231:384–99.

